# Patient-centered approaches for family planning counseling and support: A systematic review

**DOI:** 10.1101/2023.06.22.23291755

**Authors:** Dominique Meekers, Aaron Elkins, Vivian Obozekhai

## Abstract

**Background:** This paper identifies how patient-centered family planning care has been defined, conceptualized, and measured, describes tools to make family planning care more patient-centered, and discusses their impact on patient satisfaction and family planning outcomes.

**Methods:** We systematically searched PubMed and SCOPUS for documents on “patient-centered family planning counseling or support” published between 2013 and 2022. Eligibility criteria included discussion of 1) a strategy for providing patient-centered care, 2) an intervention that used a patient-centered approach, or 3) evidence of the impact of patient-centered approaches. We excluded documents that only recommended using patient-centered approaches. To assess how studies conceptualized patient-centered care, we reviewed how the concept was measured. We identified tools for patient-centered care, and mapped them against the main domains of patient-centered care. We reported the available evidence of the impact on those tools without further statistical analysis.

**Results:** Our review is based on 33 documents, including three theoretical articles and three systematic reviews. Nine studies addressed women’s experiences with family planning counseling, five discussed instruments for measuring the patient-centeredness of care, ten discussed tools for patient-centered family planning, and three discussed broader counseling programs.

We identified important differences in how patient-centered family planning care was defined and measured, although most studies emphasized patients’ needs and preferences, respect for the patient, and informed decision-making. We identified six tools for increasing the patient-centeredness of family planning counseling. None of the tools addressed all domains of patient-centered care. Evidence about the impact of these tools is scarce. Overall, the tools appeared well accepted by both providers and patients. There was some evidence that the tools improved patients’ perception about the quality of care, but no evidence that the tools improved family planning outcomes.

**Discussion:** Limitations of our study include that our search was restricted to two databases, and that the studies predominantly focused on Western countries which may limit the generalizability of the findings. Wider use of existing scales to measure patient-centered family planning care may help standardize the definition of patient-centered care and strengthen the evidence base. Although tools for patient-centered care improve the patient experience, there is a need to identify strategies for translating this into improved family planning outcomes. In addition, there is a need to test patient-centered approaches in a wider range of settings.

## Introduction

A large body of family planning literature notes the importance of patient-centered contraceptive counseling. Currently, family planning counseling is dominated by the tiered-effectiveness model, which was endorsed by the World Health Organization. In tiered-effectiveness counseling, contraceptive methods are discussed in order of effectiveness, starting with the most effective methods first (Brandi and Fuentes 2020). This approach has led to a strong emphasis on long-acting reversible contraceptive (LARC) methods.

Although the tiered-effectiveness model is compatible with shared decision-making in which the provider and patient jointly discuss which method would be most suitable for the client, it is increasingly recognized that the tiered-effectiveness model is vulnerable to unconscious provider biases. Anecdotal reports document instances of highly disrespectful family planning care, including verbal abuse, refusal of care, and unconsented care (Holt, Caglia et al. 2017, Hazel, Mohan et al. 2021). However, even when there is no explicit coercion, clients may experience implicit pressure to use --or not use-- certain contraceptive methods (Gomez and Wapman 2017). Specifically, provider preferences for the most effective contraceptive methods may cause them to overlook other factors that may be more important to the client, such as their personal values, relationship status, past contraceptive experiences, or preferences for specific contraceptive attributes. Due to the unequal power relationship between providers and clients, these biases can therefore negatively affect patient-centered care. As a result, clients may adopt a contraceptive method that suits the provider’s preference, rather than their own, which can lead to dissatisfaction with the method and contraceptive discontinuation (Downey, Arteaga et al. 2017, Gomez and Wapman 2017, Morse, Ramesh et al. 2017, Soin, Yeh et al. 2022).

In recent years there has been a rapidly growing interest in making family planning counseling more patient-centered. Patient-centered care, also known as client-centered care, is the provision of care that is unique and targeted to the individual’s circumstances, which includes centering the treatment on the individual’s needs, preferences, and values, informed decision-making, transparency, and respect (Holt, Caglia et al. 2017, Ti, Burns et al. 2019, Gawron, Simonsen et al. 2022). For family planning, patient-centered counseling involves understanding the woman’s fertility goals, contraceptive needs and preferences, education on contraceptive methods, autonomy, and a space for open dialogue (Ti, Burns et al. 2019, Dehlendorf, Fox et al. 2021). However, providing extensive patient-centered counseling tends to be time-consuming, making it difficult for providers to implement (Koo, Wilson et al. 2017, Baldwin, Overcarsh et al. 2018). To address this concern, several job aids and tools have been developed to make the provider visit more patient-centered without unnecessarily increasing the provider’s workload, but to date none of them are widely used. To accelerate the adoption of such tools, and to potentially adapt them for use in other settings, there is a need for an enhanced understanding of the specific domains of patient-centered care that they try to address.

The objective of this paper is to summarize the current state of knowledge about patient-centered family planning counseling and support, with specific emphasis on tools that are being used to help make family planning counseling more patient-centered. Specifically, our systematic review addresses the following questions:

- How has patient-centered family planning counseling and support been defined, conceptualized, and measured?
- Which tools are being used to make family planning counseling more patient-centered, which domains of patient-centered care do these tools focus on, and what is the available evidence of the impact of these tools on patient satisfaction and family planning outcomes?

An enhanced understanding of how patient-centered family planning counseling has been conceptualized in the literature can help guide the adaptation of such programs to a wider range of settings, such as developing countries. A comprehensive review of the tools used to enhance the client-centeredness of the provider-patient interaction can inform the development of similar tools suitable for different modes of family planning counseling and support, such as phone-based counseling or family planning chatbots.

## Methods

Our theoretical systematic review was implemented using the updated PRISMA 2020 guidelines for reporting systematic reviews (Page, McKenzie et al. 2021, Page, Moher et al. 2021). We applied a comprehensive search strategy aimed at producing an enhanced understanding of how different studies have conceptualized and measured patient-centered care in family planning, and at identifying the key tools or instruments that have been used to make the family planning counseling and services more patient-centered. Our review can be classified as a rapid review, by virtue of the fact that it was limited to documents in two databases (which also implied a de facto exclusion of grey literature), and covers only a ten-year period. There is no published protocol or registration for this systematic review.

### Data sources and search strategy

We conducted a systematic search of the PubMed and SCOPUS databases for articles on the subject of patient-centered family planning counseling using the search terms (“family planning” or “contraception”) and (“counseling” or “support” or “follow up”) and (“user-centered” or “client-centered” or “patient-centered”). The search was limited to documents published between January 1, 2013 and December 31, 2022.

### Screening

Two reviewers (DM and AE) independently screened the titles and abstracts of the documents for relevance. We considered documents relevant if they discussed one or more of the following aspects of patient-centered family planning counseling and support: 1) a strategy or approach for providing patient-centered counseling and support (including theoretical approaches and approaches for assessing the clients’ needs), 2) an intervention that applied a patient-centered approach, or 3) evidence of the impact of patient-centered approaches on perceived quality of care or family planning outcomes. We did not formally restrict our review to peer-reviewed studies. However, the two databases we used are skewed heavily toward peer-reviewed literature, which resulted in the de facto exclusion of grey literature.

If the two reviewers disagreed about the relevance of a document, or if either reviewer was uncertain about its relevance, the document was jointly discussed by the reviewers to achieve consensus. A small number of documents for which we did not reach consensus were retained for full document review.

### Data extraction

During the full-text review, we extracted the following data about the study characteristics: author, document title, year of publication, region (US/Europe, Africa, Asia, Latin America), document type (theoretical/conceptual paper, systematic review, methodological paper, impact evaluation, etc.), and the definition or description of patient-centered family planning care used (including domains of patient-centered care). For papers that discussed the implementation of a patient-centered family planning intervention, we extracted the type of study population (e.g., family planning clients or providers), the type of intervention (mode of delivery; name or description of any tools or aids used) and key findings. For the key findings, we used the effect measures as reported in the document (e.g., odds ratios, percentage differences). One reviewer collected data from each report, which was subsequently checked by the second reviewer. No bias assessment was conducted.

## Results

### Search results

Our initial search yielded 134 results, including 76 references from PubMed and 58 from SCOPUS. The search results were imported into the Covidence screening and data extraction tool (www.covidence.org), which identified and removed 48 duplicate records (see Figure 1). Screening of the titles and abstracts of the remaining 86 unique documents resulted in the exclusion of 41 references. The remaining 45 documents retrieved for full-text review. During the full-text review, an additional 12 documents were excluded. Two documents were excluded because we considered them to be duplicates. One of these documents – the only grey literature document – was excluded because the findings were subsequently published in peer-reviewed documents included in the review. Another document was excluded because it was a published summary of a larger article already included. Consistent with other systematic reviews, we excluded documents that did not address patient-centered approaches, but simply recommended implementing such approaches in the future (Gagliardi, Nyhof et al. 2019). This was the most common exclusion reason (n=6). Other exclusion reasons were that the study did not elaborate on the family planning counseling approach (n=2) or only included a study protocol (n=1). One study was deemed irrelevant because it only addressed provider perceptions about the quality of their training. Hence, 33 documents were retained for our review. The data we extracted from these documents are available on the Harvard Dataverse repository (Meekers, Elkins et al. 2023).

**Figure 1:**
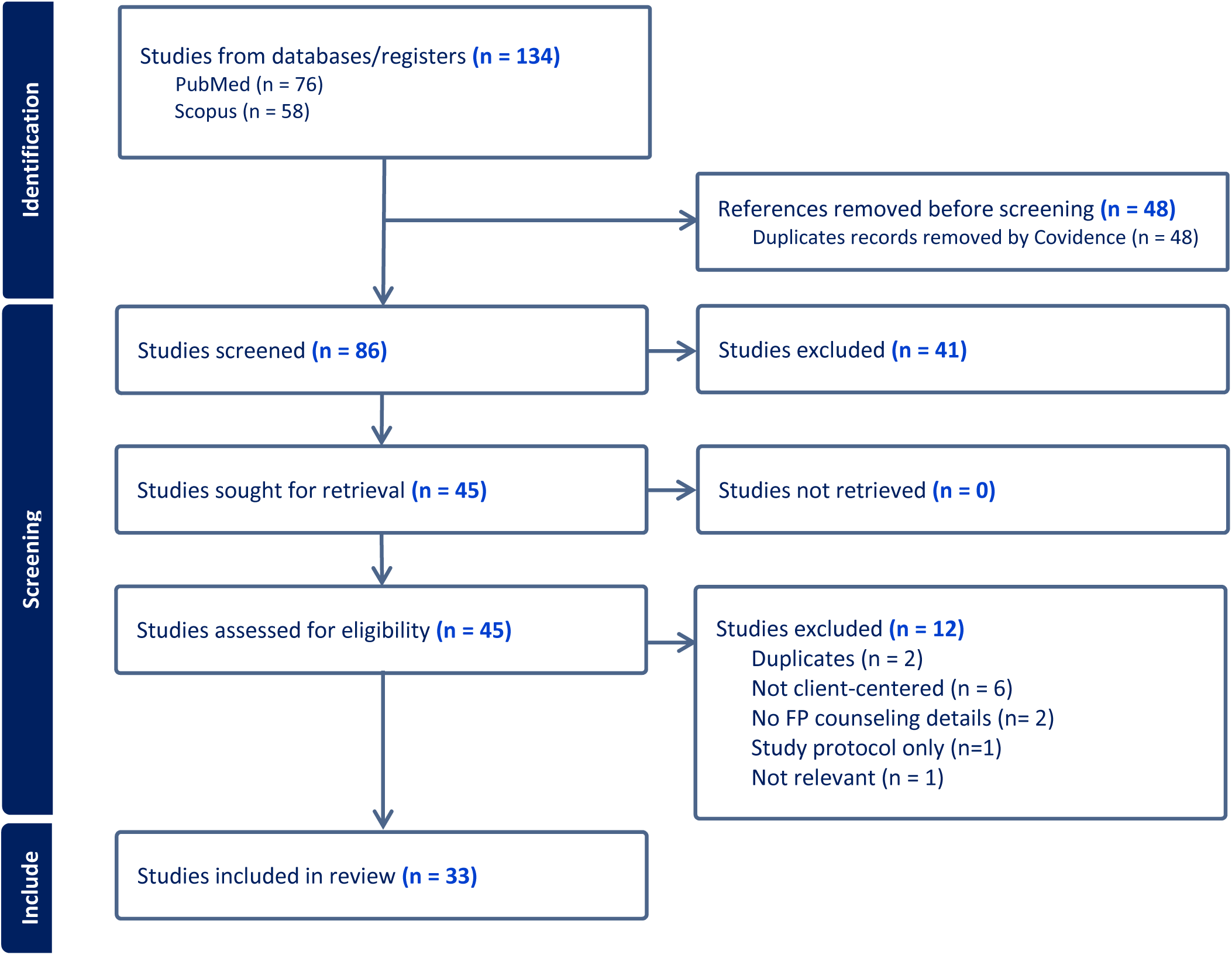
PRISMA flow chart.

### Characteristics of the included literature

The 33 full-text documents retained for this rapid review included three theoretical and/or conceptual articles (Holt, Caglia et al. 2017, Morse, Ramesh et al. 2017, Brandi and Fuentes 2020) and three systematic reviews (Fox, Reyna et al. 2018, Gagliardi, Nyhof et al. 2019, Soin, Yeh et al. 2022). Eight studies focused on women’s experiences with family planning counseling, their perceived quality of care, and their counseling preferences (Assaf, Wang et al. 2017, Gomez and Wapman 2017, Holt, Zavala et al. 2018, Callegari, Tartaglione et al. 2019, Ti, Burns et al. 2019, Hazel, Mohan et al. 2021, Singal, Sikdar et al. 2021, Hamon, Hoyt et al. 2022). One study discussed how women’s contraceptive preferences change based on their personal circumstances and experiences, resulting in an iterative process of contraceptive decision-making (Downey, Arteaga et al. 2017). Five of the studies developed, validated, or applied a survey instrument for measuring the level of patient-centered family planning care (Dehlendorf, Henderson et al. 2016, Dehlendorf, Henderson et al. 2018, Carvajal, Mudafort et al. 2020, Dehlendorf, Fox et al. 2021, Welti, Manlove et al. 2022). Four studies discussed a reproductive goal screening tool (Baldwin, Overcarsh et al. 2018, Madrigal, Stempinski-Metoyer et al. 2019, Stulberg, Dahlquist et al. 2019, Gawron, Simonsen et al. 2022), six a contraceptive decision-making tool (Donnelly, Foster et al. 2014, Koo, Wilson et al. 2017, Dehlendorf, Fitzpatrick et al. 2019, Dehlendorf, Reed et al. 2019, Dev, Woods et al. 2019, Callegari, Nelson et al. 2021), and three discussed broader counseling programs or curricula (Kamhawi, Underwood et al. 2013, Loyola Briceno, Kawatu et al. 2017, Worthington, Oyler et al. 2020). Most of the studies that assessed a patient-centered family planning intervention, tool, or measurement instrument were conducted in the United States (n=20), while only four studies were done in Africa (Assaf, Wang et al. 2017, Dev, Woods et al. 2019, Hazel, Mohan et al. 2021, Hamon, Hoyt et al. 2022), one in the Middle East (Kamhawi, Underwood et al. 2013), one in Asia (Singal, Sikdar et al. 2021) and one in Latin America (Holt, Zavala et al. 2018).

## Results

### Definitions and conceptualization of patient-centered counseling and care

In the literature, the terms “patient-centered” and “client-centered” are used interchangeably (the term “person-centered” generally focuses on more holistic, longer-term goals). Our literature review indicates that there is no universally agreed upon general definition of patient-centered care, and consequently there are differences in what is considered patient-centered care in family planning counseling and support. Only 18 of the 33 documents either included a clear definition of client- or patient-centered family planning counseling, or clearly described key features or domains of patient-centered family planning counseling or care. However, studies that identified problems with quality of family planning care, such as negative experiences with providers, tend to address similar topics without referring to them as domains of patient-centered care (Downey, Arteaga et al. 2017, Gomez and Wapman 2017, Callegari, Tartaglione et al. 2019).

Three of the studies included in our review made reference to the 1990 Judith Bruce Quality of Family Planning Care (Assaf, Wang et al. 2017, Holt, Caglia et al. 2017, Hazel, Mohan et al. 2021). That original Bruce framework identified six distinct elements of the quality of family planning care “that clients experience as critical”, including 1) the choice of methods that are offered on a reliable basis, 2) the information provided to the client, 3) the technical competence of the provider, 4) the interpersonal relations between the providers and clients, 5) the mechanism to promote continuity of care (e.g., follow-up visits), and 6) the availability of an appropriate constellation of acceptable and convenient family planning services (Bruce 1990). Although this framework is very broad, it emphasizes the importance of the client’s perspective about the quality of care, including the provider-patient relationship. Consequently, the framework forms the basis for much of the contemporary discussions about patient-centeredness family planning care, and women’s autonomy in family planning decision-making.

Although the reviewed studies varied in how they defined patient-centered care, several either referred to the 2001 Institute of Medicine definition of patient-centered healthcare or built on that definition (Dehlendorf, Henderson et al. 2018, Ti, Burns et al. 2019, Carvajal, Mudafort et al. 2020). The Institute of Medicine (renamed to National Academy of Medicine in 2015) described patient-centered care as “care that is respectful of and responsive to individual patient preferences, need, and values and ensures that patient values guide all clinical decisions” (Institute of Medicine 2001: 40). A number of other studies used definitions or descriptions of patient-centered care or counseling that referred to these same elements. For example, Brandi and Fuentes (2020: s876) stated that patient-centered counseling “aims to provide education to patients that integrates evidence-based recommendations based on patient preferences, recognizing that patients’ individual values and preferences should be an integral factor in decisions made about their health care [and ensures that] patients function as experts on their preferences and needs and providers function as experts on the medical evidence.”

Although studies used different terminology, definitions, and approaches for patient-centered care, Holt, Caglia et al. (2017: 1) note that they all acknowledge “the essential role of individuals’ preferences, needs and values, and the importance of informed decision-making, respect, privacy and confidentiality, and non-discrimination.”

Despite these commonalities, Gagliardi, Nyhof et al. (2019) noted that a better understanding of the different domains of patient-centered care can facilitate more accurate – and more consistent – measurement, which in turn can inform the design of strategies to strengthen patient-centered care. In their theoretical rapid review of the evidence on the patient-centeredness of women’s health care, they mapped studies against the dimensions of patient-centered healthcare that were previously identified by (McCormack, Treiman et al. 2011). McCormack argued that there are six main domains of patient-centered care:

1. **Fostering the relationship between the provider and client.** This domain includes building rapport with the patient, trust in the provider’s technical competency, his/her honesty and openness, demonstrating that the provider cares about what is best for the patient, as well as discussing the provider and patient’s respective roles responsibilities.
2. **Reciprocal exchange of information between provider and client.** Sub-domains include obtaining information about the patient’s information needs, beliefs, and preferences, and sharing information and resources with the patient.
3. **Recognizing the patient’s emotions and responding to them.** By asking the patient questions about their emotions, the provider signals an understanding of the patient’s situation and shows empathy.
4. **Managing uncertainty.** This domain includes assessing sources of the patient’s uncertainty (e.g., about side-effects or life changes), and using emotion- and problem-focused strategies to address them.
5. **Making decisions.** Subdomains include communicating what the provider and patient expect about their respective roles in the decision-making process, sharing information to support decision-making, and offering opportunities to participate in decision-making.
6. Enabling patient self-management, including advising the patient, helping the patient plan, and arrange for follow-up.

The authors noted that each study in their review defined and measured patient-centered care differently, and none of them addressed all six domains (Gagliardi, Nyhof et al. 2019). The most commonly addressed domains were exchanging information, making decisions and fostering the relationship. The authors noted that none of the studies in their review measured patient-centered care as comprehensively as the McCormick framework.

### Measurement of the patient-centeredness of family planning counseling

Many of the studies included in our review attempted to measure the extent to which family planning clients perceived the interaction with the provider as patient-centered. However, in absence of a universally agreed upon definition of patient-centered counseling, we found a lack of consistency in how it has been measured. Key questions focused on the following areas: 1) soliciting information about the clients’ reproductive preferences; 2) soliciting the clients’ personal opinion about specific methods; 3) soliciting information about the expected role of the provider in the decision-making process; 4) the provider’s level of compassion and empathy; 5) the provider’s level of respect for the client; and 6) providing the patient opportunities to ask questions. Illustrative examples of questions asked to assess provider performance with respect to each of these topics are shown in Table 1.

**Table 1:** Illustrative questions used for measuring the patient-centeredness of family planning counseling.

|  |
| --- |
| **Soliciting information about the patient's reproductive preferences**<br>• Did the provider discuss how many children you would like to have? (Kamhawi, Underwood et al. 2013)<br>**Soliciting the clients' person opinions about specific contraceptive methods**<br>• Did the provider ask what birth control method sounded like a good choice to you? (Brandi and Fuentes 2020)<br>• Did the provider ask you about your preference in contraceptive methods? (Hazel, Mohan et al. 2021)<br>• Do you feel the provider advocated a specific method for you during the consultation? (Hazel, Mohan et al. 2021)<br>• Did the provider ask if you had a method in mind before coming to the clinic? (Kamhawi, Underwood et al. 2013)<br>**Meeting expectations about the expected role of the provider in the decision-making process**<br>• How satisfied are you with the decision making process about which birth control method you will use? (Dehlendorf, Henderson et al. 2018)<br>• Who do you feel made the decision about the chosen method? (Dehlendorf, Fitzpatrick et al. 2019)<br>• To what extent was the choice of your contraceptive method a shared decision between you and your provider? (Koo, Wilson et al. 2017)<br>**Provider compassion and empathy**<br>• To what extent do you agree that the provider showed care and concern about you as a person? (Koo, Wilson et al. 2017)<br>**Provider respect for the patient**<br>• Did the provider greet you respectfully? (Hazel, Mohan et al. 2021)<br>• Did the provider make critical or judgmental comments about a) the number of children you have, b) your fertility plans, c) your partner or marital status, d) the involvement of your partner in family planning, e) your sexual activity, f) involvement of your parents, g) your age in reference to family planning, h) your preferred contraceptive method (Hazel, Mohan et al. 2021)<br>• To what extent do you agree that the provider did not judge you? (Koo, Wilson et al. 2017)<br>**Providing the patient opportunities to ask questions**<br>• Did the provider ask what questions you had about any of the methods? (Brandi and Fuentes 2020).<br>• Did the provider interrupt you while you were speaking? (Hazel, Mohan et al. 2021) |

In an attempt to standardize measurement of the patient-centeredness of family planning counseling, some authors have developed and validated scales to measure the level of patient-centeredness of the family planning counseling visit. One of the most comprehensive tools for measuring patient-centeredness of family planning counseling and services visits we identified was the Interpersonal Quality in Family Planning Care (IQFP) scale (Dehlendorf, Henderson et al. 2016, Dehlendorf, Henderson et al. 2018). The IQFP is a validated 11 item scale that measures distinct aspects of the interpersonal communication between provider and patient. Specifically, the scale is based on eleven questions that ask family planning patient to rate the provider on the following issues:

1. Respecting me as a person
2. Showing care and compassion
3. Letting me say what mattered to me about my birth control method
4. Given me an opportunity to ask questions
5. Taking my preferences about my birth control seriously
6. Considering my personal situation when advising me about birth control
7. Working out a plan for my birth control with me
8. Giving me enough information to make the best decision about my birth control method
9. Telling me how to take or use my birth control most efficiently
10. Tell me the risks and benefits of the birth control method I chose
11. Answering all my questions.

Patients rated each of these 11 included items on a 5-point Likert scale, ranging from “poor” to “excellent.” Because the large majority of users rated the items as excellent, the authors dichotomized the item responses into the highest possible rating (excellent) versus all lower scores. Validity tests showed that the IQFP scale was associated with clients’ level of satisfaction with the family planning counseling they received, as well as satisfaction with their chosen contraceptive method. Furthermore, higher scores on the IQFP scale was associated with positive provider communication practices, including eliciting the patients’ perspectives and demonstrating empathy. Multivariate analyzes show that high scores on the IQFP scale were associated with positive family planning outcomes, including continuation of the chosen method at six months (OR=1.81 [1.09-3.00]) and use of an effective method at six months (OR 2.03 [1.16-3.54]). Examination of the different scale items suggested continuation of the chosen method at six months was higher when the provider invested in the early part of the counseling session (OR=2.32 [1.24-4.32]) and elicited the patient perspective (OR=1.79 [1.01-3.16]). However, showing empathy or investing in the end of the session (e.g., by discussing follow-up etc.) had no effect on contraceptive continuation (Dehlendorf, Henderson et al. 2016).

Recognizing that the large number of items in the IQFP may limit its usefulness as a tool for assessing provider performance, a reduced version of the scale has been produced (Dehlendorf, Fox et al. 2021). The Person-Centered Contraceptive Counseling (PCCC) scale asks family planning clients to think about their last provider visit and ask them how they would rate the provider on the following items from the original IQFP scale:

1. Respecting me as a person
2. Letting me say what mattered to me about my birth control method
3. Taking my preferences about my birth control seriously
4. Giving me enough information to make the best decision about my birth control method

Because the 4-item PCCC scale reduces the burden of data collection compared to the more comprehensive IQFP scale, it is more feasible to use it as a tool for improving the quality of the provider-patient interaction. The PCCC scale has since been incorporated into the questionnaire of the National

Survey of Family Growth (NSFG). Analyses of the NSFG show that while most respondents gave their provider an excellent rating on each of the four scale items, clients’ experiences related to person-centered care varied across sociodemographic groups, with low income women, sexual minorities, and women with limited English proficiency giving their provider lower ratings for patient-centeredness (Welti, Manlove et al. 2022). The authors hypothesized that these lower ratings may reflect discrimination and/or a lack of cultural competency. The authors also noted that the association between low English proficiency and lower PCCC rating highlights that providing patient-centered care may require language concordance between providers and patients.

The fact that the 4-item PCCC was incorporated in the NSFG survey suggests that it is likely to be more widely adopted. If so, it will further enhance consistency in measurement of the level of patient-centeredness of family planning counseling, and increase comparability across different studies.

### Tools for increasing the patient-centeredness of family planning counseling

Our review of the literature identified a wide range of tools that have been developed and used by interventions that offer patient-centered family planning counseling (see Table 2), including the following:

**Table 2:** Tools to promote patient-centered family planning counseling, by author.

|  | Patient-Centered Care Tool |  |  |  |  |  |  |
| --- | --- | --- | --- | --- | --- | --- | --- |
|  | One Key Question | Family planning quotient/reproductive life index | Smart Choices | My Birth Control | MyPath | iMacc | Other tools |
| Baldwin, Overcarsh et al. (2018) | √ | √ |  |  |  |  |  |
| Brandi and Fuentes (2020) | † |  |  |  |  |  |  |
| Callegari, Nelson et al. (2021) |  |  |  | √ | √ |  |  |
| Dehlendorf, Fitzpatrick et al. (2019) |  |  |  | √ |  |  |  |
| Dehlendorf, Reed et al. (2019) |  |  |  | √ |  |  |  |
| Dev, Woods et al. (2019) |  |  |  |  |  | √ |  |
| Donnelly, Dehlendorf et al. (2019) |  |  |  |  |  |  | √ |
| Gawron, Simonsen et al. (2022) | √ | † |  |  | † |  |  |
| Kamhawi, Underwood et al. (2013) |  |  |  |  |  |  | √ |
| Koo, Wilson et al. (2017) |  |  | √ |  |  |  | † |
| Madrigal, Stempinski-Metoyer et al. (2019) | † | √ |  | † |  |  | † |
| Morse, Ramesh et al. (2017) | † |  |  |  |  |  | † |
| Stulberg, Dahlquist et al. (2019) | √ |  |  |  |  |  |  |

Note: √ Tool was applied or tested in the study; † tool was mentioned/discussed in the study.

#### One Key Question (OKQ)

Several studies used One Key Question (Baldwin, Overcarsh et al. 2018, Stulberg, Dahlquist et al. 2019, Gawron, Simonsen et al. 2022). OKQ is a screening tool that aims to 1) help determine a patient’s preferences about a future pregnancy, 2) facilitate a subsequent discussion about the patients’ reproductive goals, and 3) encourage the provider to offer patient-specific contraceptive counseling. The OKQ tool was originally designed by the Oregon Foundation for Reproductive Health (Hunter, Meieran et al. 2012). It is currently licensed by Power to Decide (www.powertodecide.org), which offers certification training. To use OKQ, providers are required to ask clients “Would you like to become pregnant in the next year? (yes; no; unsure; I am OK either way)”, which must be followed by comprehensive patient-centered counseling that is consistent with the client’s response. All clients are offered preconception counseling (including screening for potential high-risk pregnancies, advice to reduce alcohol or tobacco use, etc.), and informed about the benefits of adequate child spacing and about contraceptive options. For clients who do not wish to become pregnant, information is provided about effective contraceptive methods, their correct use, and what to do in case of accidental incorrect use. Clients who are unsure about their pregnancy intentions or are OK either way, are offered both contraceptive and preconception care, tailored to their specific goals (Hunter, Meieran et al. 2012). Given that there is flexibility in how the provider responds, use of OKQ does not necessarily result in patient-centered counseling (Gawron, Simonsen et al. 2022).

#### Family Planning Quotient and Reproductive Life Index (FPQ/RepLI)

Like the OKQ, the FPQ/RepLI tool was designed to facilitate discussions and decision-making about reproductive life goals and family planning (FPQ) (Baldwin, Overcarsh et al. 2018, Madrigal, Stempinski-Metoyer et al. 2019). The FPQ/RepLI is described as a patient-centered tool that is intended to be incorporated into the patient’s electronic medical record. The tool visually depicts a patient’s reproductive life plan, enables tracking of progress toward the patient’s reproductive goals, and helps the provider discuss the patient’s needs and options. The FPQ/RepLI tool is completed before the patient sees the provider. To collect the necessary information for the FPQ/RepLI tool, a health educator first speaks with the patient about her sexual, gynecologic and obstetric history and completes the tool.

FPQ/RepLI is comprised of four main components. The first component is a graph that visualizes the Family Planning Quotient, which is the ratio of the number of children the patient already has, including both biological and non-biological children, over the desired number of children. When the patient’s FPQ is below one, she has not yet achieved her reproductive goals; when the FPQ equals one, her reproductive goal has been met; and when it is larger than one, she has already exceeded her desired number of children. The second part of the tool is a decision-making tree for selecting the types of contraceptives the patient should be counseled on. For women who have met or exceeded their reproductive goal, counseling focuses on reversible or permanent long-term contraceptive methods. For women, when who have not yet met their reproductive goal, the One Key Question is used to determine the type of contraceptive counseling. Women who wish to get pregnant in the next year are counseled on short-acting contraceptives methods. Women who wish to get pregnant later are counseled on both short-acting contraceptives and long-acting reversible methods. The third component of the tool is the Reproductive Life Index (RepLI), which tracks annual progress in the FPQ, pregnancy outcomes (including unplanned births), as well as other relevant outcomes (e.g. adopted children and stepchildren). Finally, the fourth part consists of a table that tracks annual changes in the type of contraceptive method used (Madrigal, Stempinski-Metoyer et al. 2019). A unique feature of the FPQ/RepLI tool is that it provides a longitudinal, graphical view of the patient’s progress toward the stated reproductive goal. As such, it takes into account that a woman’s reproductive goals can be fluid and change over time.

#### Smart Choices

Smart Choices is a computer-based tool that aims to improve contraceptive counseling aid by making the counseling session more comprehensive, better tailored to the patient’s need and preferences, and by increasing the patient’s ability to have an active role in contraceptive decision-making (Koo, Wilson et al. 2017). A detailed description of the tool is provided in Wilson, Krieger et al. (2014). The Smart Choices tool is designed to be downloaded and used by a wide range of clinics, provided that they have a computer and printer. The tool is then used by patients while they wait to meet with their provider. The first component of the tool is a questionnaire that asks the patient about childbearing plans and intentions, including about things the patient may desire before having a baby (e.g., complete education). This component also collects information about the patient’s contraceptive experience (including level of method satisfaction), partner influences on pregnancy preferences and contraceptive use, menstrual problems, sexual risk behavior, and asks about questions/concerns about contraceptives or sexual health the patient would like to discuss with the provider. A printed form with the patient’s answers helps clarify to the provider which issues the patient is concerned about, and helps streamline the counseling topics.

The second component of the tool consists of an interactive, audio-visual guide that allows the patient to get in-depth information about different methods. The tool enables the patient to select contraceptive methods with specific attributes she may prefer (STI protection; non-prescription; non-hormonal; instant sex; easy to hide; works immediately; lighter periods). Contraceptive methods with the selected attributes are organized by level of effectiveness. For each method, the tool provides a two minute audio-visual presentation and/or detailed text about the method. Smart Choices does not recommend or encourage use of a specific method (Wilson, Krieger et al. 2014, Koo, Wilson et al. 2017).

#### My Birth Control

My Birth Control is a tablet-based interactive family planning decision-making tool used to assess women’s contraceptive values and preferences and help them select a conceptive method that matches those preferences (Dehlendorf, Fitzpatrick et al. 2019, Dehlendorf, Reed et al. 2019). Like several other tools, My Birth Control is intended to be used before visiting a provider. The tool aims to improve use of best practices in shared contraceptive decision-making. To achieve that, the tool provides information about different contraceptive methods, inquires which contraceptive features are most important to the users, conducts a short medical history check, and then recommends contraceptive methods based on the information provided by the user. A printout of the user’s answers and the recommended methods can then be shared with the provider to inform the counseling session.

The My Birth Control tool is available at https://clinic.mybirthcontrol.org. The first part of the My Birth Control tool aims to address common questions that patients have about modern contraceptive methods, including the effectiveness of the various methods, how they are used, how often the method needs to be administered or renewed, the potential side-effects of the method, and what to do if or when the patient decides she wants to get pregnant. When the user selects “how well does it prevent pregnancy,” the user sees an infographic that shows that 85 out of 100 women will get pregnant during the first year of not using a contraceptive method. The user can then select a contraceptive method from a list of modern methods (arranged from most to least effective) to see how many unintended pregnancies are expected for that method. Next, the tool invites the patient to click on the icon presenting a modern method to get a short description of how it is used. Similarly, the patient can select a method to see a graph that indicates how often it needs to be used. The section of the tool that addresses potential side-effects first gives short descriptions of common side-effects that users may find positive, side-effects that one may find annoying, and side-effects that one should not worry about. The user can then select a modern method to see the specific side-effects associated with that method.

The second part of My Birth Control gathers information about when the patient thinks she may want to get pregnant, if at all, and her preferences for specific method attributes. Specially, the questions gather detailed information about the client’s preferences regarding method effectiveness, convenience of use, and the way the method is administered. The patient is also asked about her level of tolerance for several specific side-effects, including spotting/irregular bleeding, not having a period, heavier periods or cramping, and weight gain. Similarly, clients are asked how they feel about various potential method benefits (decreased acne, not having a period, decreased cramping, less heavy periods). Clients are also asked which modern method they have already used, and whether they liked the method. After inquiring about possible contraindications (high blood pressure, smoking, etc.), My Birth Control will recommend options that match the user’s preferences. The tool first shows contraceptive methods that match the preferred effectiveness, then methods that match the preferred mode of administration and frequency of use, and finally, methods that match the client’s preferences regarding potential side-effects and benefits. It also lists the methods that are not a good fit for the client’s preferences, recognizing that the patient may still decide to use them.

#### MyPath

MyPath is a web-based family planning decision-support tool designed to increase reproductive health counseling and services during primary care visits, optimize the patient’s health prior to pregnancy, and increase support for family planning decision making (Callegari, Nelson et al. 2021, Gawron, Simonsen et al. 2022). Detailed information about MyPath is available at https://info.mypathtool.org/. The tool is designed to be used before visiting a primary care provider. MyPath uses a broad patient-facing approach, with a strong focus on reproductive autonomy. To achieve this, MyPath enables women to more easily communicate their reproductive goals and preferences, strengthens their self-efficacy by informing them about their contraceptive options, and improves the provider-patient relationship by encouraging patient-centered communication.

Key components of the tool include sections that 1) solicit information about the client’s feelings and preferences regarding pregnancy and childbearing, 2) provide information about the menstrual cycle and fertility, 3) provide information about pre-pregnancy health, and 4) help identify a suitable birth control method, using the previously discussed My Birth Control tool (Callegari, Nelson et al. 2021). The first section includes a question that asks the patient to articulate her reproductive preference, recognizing that women may have ambivalent feelings about pregnancy (e.g., women who wish to avoid pregnancy are not necessarily unhappy if they become pregnant anyway). The second section clarifies when during the menstrual cycle women are most likely to conceive, and addresses misconceptions about pregnancy risk. The third section provides information about the effect of both physical and mental health on pregnancy, and aims to stimulate provider-patient discussions about actions that can improve pre-pregnancy health (including life style changes, maintaining a healthy weight, taking folic acid, etc.). Finally, the fourth section uses the *My Birth Control* tool to 1) educate the patient about various aspects of different contraceptive methods (including ease of use, potential side-effects, return to fertility, etc.); 2) get information about the client’s preferences regarding these attributes; and 3) help her select an appropriate contraceptive method that is consistent with those preferences.

#### Interactive Mobile Application for Contraceptive Choice (iMACC)

iMACC is an interactive, patient- faced family planning decision-making app for mobile phones (Dev, Woods et al. 2019). Use of the iMACC phone app is designed to streamline family planning counseling and help women make informed, personal, contraceptive choices. It was specifically designed for post-partum women, as this group may have unique preferences with respect to the features of their contraceptive method. For example, they may prefer methods that allow a quick return to fertility or methods that can be used safely while breastfeeding. iMACC is intended to be self-administered while clients wait to visit their health provider. Unlike many other tools, clients can use iMACC independently (i.e., without provider involvement) if they so desire, and the tool can help them select a contraceptive method that suits their needs and preferences by themselves. For women who prefer more provider input, the tool can help them determine which questions to ask the provider during counseling, which in turn helps streamline the counseling session.

The iMACC tool combines text and images and includes 14 health history questions and 48 queries to access individual preferences, preferences and concerns about family planning (Dev, Woods et al. 2019, Dev 2023) The health history section covers topics such as pregnancy outcomes, breastfeeding, high blood pressure, chronic headaches, cigarette smoking, etc. Family planning preferences include questions not only about the desired number of children and the preferred timing, but also how important it is for the patient to avoid pregnancy at this time. Contraceptive history questions identify the different contraceptive methods the patient has tried, and assesses whether they had a good experience with that method and whether they would use it again in the future. Users are then asked to identify the three contraceptive attributes that are most important to them (effectiveness, convenience of use, concealability, reduced menstrual flow, side-effects, duration, and cost of the method). The tool also inquires about the partner’s attitude toward family planning, including whether there are any methods the partner would not feel comfortable using. This is followed by a series of detailed questions about the user’s preferences regarding specific aspects of method conveniences, concealability, menstrual flow preferences, side effects, cost, the frequency of method administration, and the time it takes to return to fertility. The tool then provides information on six modern methods, and list methods that are consistent with each category of attribute preferences (e.g., methods with the desired effectiveness, methods that avoid undesired side effects, etc.).

#### Other tools

A few studies discussed alternative tools, but lacked detailed information about them. For example, Donnelly, Foster et al. (2014) noted they intend to develop a new contraceptive decision support tool that is based on the Option Grid model. Other tools mentioned briefly include reproductive life planning tools, such as the Reproductive Health Self-Assessment Tool (RH-SAT) and contraceptive decision-making tools such as Bedsider, My Method, Method Match, and Best Method for Me (Koo, Wilson et al. 2017, Morse, Ramesh et al. 2017).

In addition, some authors did not use a specific tool, but relied on a broader package of tools. For example, Kamhawi, Underwood et al. (2013) discussed a package of tools that included the WHO’s medical eligibility wheel, service provider and patient cue cards, and informational posters. Still others described broader training curricula (Loyola Briceno, Kawatu et al. 2017, Worthington, Oyler et al. 2020).

### Mapping the tools against the domains of patient-centered care

As shown above, there is considerable variation in the content covered by the different tools that are being used to stimulate patient-centered family planning care. These content differences imply that the tools do not necessarily focus on all domains of patient-centered care. Table 3 maps the different tools against the six main domains of patient-centered care that are being addressed.

**Table 3:** Main domains of patient-centered family planning care explicitly addressed by different tools.

|  | Patient-Centered Care Domains |  |  |  |  |  |
| --- | --- | --- | --- | --- | --- | --- |
|  | Building provider-patient relationship | Exchanging information | Addressing patient emotions | Managing uncertainty | Making decisions | Enabling self-management |
| One key question (OKQ) | † | Tool collects client's desire to get pregnant in the next year.<br>†† Provider counsels on preconception health, benefits of child spacing and contraceptive options. | † | †† Provider provides both contraceptive and preconception care for clients who are unsure about their pregnancy intentions. | † | †† Provider explains what do to in case of incorrect method use. |
| Family planning quotient and Reproductive Life Index (FPQ/RepLI) | Tool is part of the patient's record, which provides continuity across multiple provider visits. | Tool tracks client's reproductive goal relative to actual family size (incl. non-biological children) over time.<br>Tool/health educator collects sexual, gynecologic, and obstetric history.<br>† Provider counsels on tools that match reproductive goals/intentions. | † | Tool uses a longitudinal approach, which allows reproductive goals to be fluid. | Tool shows progress toward long-term reproductive goal. Results determine whether the provider counsels on short-acting, long-acting, or permanent methods. | † |
| Smart Choices | Tool asks patient about questions she wants to discuss with the provider. Provider addresses questions/concerns patient listed on the tool printout. | Tool collects patient info about childbearing plans and intentions, contraceptive history and experience, partner attitudes, sexual risk behavior.<br>Tool provides in-depth audio-visual information about the range of contraceptive methods.<br>Tool collects patient preferences regarding contraceptive attributes and provides detailed information about methods that match those preferences. | Tool inquires about partner influences, past experiences with contraceptives.<br>Tool inquires about client's concerns about contraceptives or sexual health (open-ended). | † | Tool identifies contraceptive methods that fit client's preferences but does not recommend a specific method. | † |
| My Birth Control | Tool invites questions patient wants to discuss with the provider (open-ended). Provider addresses questions patient listed on the tool printout. | Tool addresses common questions about modern contraceptive use, and detailed information about specific methods.<br>Tool collects patient info about pregnancy intentions, contraceptive history and experience, and contraindications.<br>Tool collects patient preferences regarding desired and undesired contraceptive attributes. | Tool inquires about past experiences with contraceptives.<br>Tool inquires how much patient cares about various method attributes and how she feels about different side-effects or benefits. | † | Tool recommends contraceptive methods that fit client's preferences and past experiences. | † |
| My Path | Same as My Birth Control. | Same as My Birth Control, plus:<br>Tool provides information about menstrual cycle, pregnancy risk, and pre-pregnancy physical and mental health. | Same as My Birth Control, plus:<br>Tool assesses emotions regarding potential pregnancy. | Tool allows clients to express ambivalence regarding pregnancy intentions. | Same as My Birth Control. | † |
| Interactive Mobile Application for Contraceptive Choice (iMACC) | † | Tool collects patient info about medical eligibility, fertility and family planning intentions, contraceptive history and experience.<br>Tool provides family planning info and dispels myths and misconceptions. | Tool inquires about past experiences with contraceptives; partner's attitudes/preferences.<br>Tool inquires about preference for various method attributes; concerns. | Tool allows clients to express ambivalence regarding pregnancy intentions. | Tool identifies contraceptive methods that fit client's preferences. | † |

Note: † Up to discretion of the provider; †† Not included in the tool itself, but part of the associated provider certification.

The results showed that the reviewed tools tend to focus mostly on four of the six domains of patient-centered care. All tools directly addressed the exchange of information and all but one directly assisted with decision-making. Most tools also address one or more of the other domains indirectly, typically by gathering relevant information which is then expected to encourage the provider to take up those issues during the counseling.

Although building rapport between the provider and patient is recognized as a key component of patient-centered counseling, none of the tools addressed it directly. Three of the tools attempted to address the power imbalance between the provider and client, by explicitly inviting the user to list any questions or concerns she may have, and that she would like the provider to address. The FPQ/RepLi tool is unique because the tool help ensure continuity of care, which is known to be important for building a trusted provider-patient relationship. Because FP/RepLi is integrated into the patient’s medical record, the tool helps provide continuity across multiple provider visits (Baldwin, Overcarsh et al. 2018, Madrigal, Stempinski-Metoyer et al. 2019).

All of the reviewed tools collected information about the patient’s pregnancy preferences and/or reproductive goals. All but OKQ collect information about the client’s contraceptive history. Smart Choices, My Birth Control, My Path and iMACC also inquired about the client’s experience with each of the previously used methods and preference for specific method attributes (Koo, Wilson et al. 2017, Dehlendorf, Fitzpatrick et al. 2019, Dev, Woods et al. 2019, Callegari, Nelson et al. 2021). These same tools also provided the patient with detailed information about contraceptive methods, including their effectiveness, use, side-effects, etc. The OKQ and FPQ/RepLI tools do not provide the patient with information about contraceptives and leave that responsibility to the provider.

Smart Choices, My Birth Control, My Path, and iMACC all make some provisions for clients to express their emotions (Koo, Wilson et al. 2017, Dehlendorf, Fitzpatrick et al. 2019, Dev, Woods et al. 2019, Callegari, Nelson et al. 2021). All four of these tools inquire how the patient feels about previously used contraceptive methods. My Birth Control, My Path, and iMACC also ask how the patient feels about specific method attributes, including potential side-effects. Smart Choices and iMACC also inquire about partner influences on contraceptive use (e.g. partner attitudes, preferences for specific methods). As such, the tools help the provider identify patient emotions that may affect contraceptive preferences, which then guides the counseling.

To facilitate decision-making, Smart Choice, My Birth Control, My Path and iMACC identify and/or recommend contraceptive methods that match the client’s preferences. The RPQ/RepLI tool does not identify methods that are suitable for the client, but the level of progress toward the client’s long-term reproductive goal is used by to provider to determine whether to counsel the patient on short-action, long-action, or permanent methods. The OKQ tool does not include decision-making assistance, which is left up to the discretion of the provider.

Overall, the reviewed tools do not appear to have been designed to thoroughly manage uncertainty. Three of the tools (OKQ, My Path and iMACC) do explicitly allow clients to express ambivalence toward their pregnancy intentions (Hunter, Meieran et al. 2012, Dev, Woods et al. 2019, Callegari, Nelson et al. 2021), while FPQ/RepLI recognizes that clients’ reproductive goals may change over time (Baldwin, Overcarsh et al. 2018, Madrigal, Stempinski-Metoyer et al. 2019). While these tools help alert the provider to this uncertainty, how to manage this is typically left to the discretion of the provider. A notable exception is OKQ, as the associated certification training requires that clients who are uncertain about their pregnancy intentions should be offered broader counseling that includes both preconception health and contraceptive options (Hunter, Meieran et al. 2012). None of the reviewed tools include components that enable or facilitate self-management, almost completely leaving that up to the provider. The OKQ certification does explicitly instruct providers to explain what clients should do in case of incorrect method use.

### Impact of patient-centered counseling approaches and tools

Table 4 summarizes the state of knowledge about the acceptability of various tools to promote patient-centered family planning, as well as the effect of those tools on the provider-patient interaction and quality of care, and on the client’s contraceptive knowledge, decision-making, method adoption, and method continuation. Because several of the tools to promote patient-centered family planning are relatively new, much of the evidence-is based on small-scale pilot studies and should be interpreted with caution. It is also important to note that two of the tools, OKQ and FPQ/RepLi, are intended to be integrated with regular physician visits, with the broad aim of increasing the likelihood that the client’s reproductive goals are discussed during such visits. By contrast, tools such as Smart Choices, My Birth Control, My Path, and iMACC are all intended to be used with clients who are specifically seeking family planning counseling or services.

**Table 4:** Summary of impact of tools to promote patient-centered family planning counseling.

| Tool/<br>Author | Study type | Ease of use/acceptability | Effect on quality of care<br>and provider-patient interactions | Effect on<br>contraceptive<br>knowledge | Effect on contraceptive<br>decision-making, use,<br>and continuation |
| --- | --- | --- | --- | --- | --- |
| **OKQ** |  |  |  |  |  |
| (Baldwin,<br>Overcarsh et al.<br>2018) | RCT with post-test survey<br>(clients: OKQ<br>n=39, FPQ n=37;<br>providers OKQ<br>n=43, OKQ n=36). | OKQ patients were less likely than FPQ<br>patients to find the tool helpful and use it to<br>track reproductive health goals (51% vs. 76%,<br>p=.02). | OKQ and FPQ clients were equally likely to find the tool<br>helpful in communicating their reproductive goals to<br>their provider (68% vs. 66%, p=.88).<br>OKQ providers were more likely than FPQ providers to<br>agree the tool helped focus their counseling, but the<br>effect was not significant (50% vs. 37%, p=.25). |  |  |
| (Gawron,<br>Simonsen et al.<br>2022) | Cross-sectional<br>pre-post patient<br>chart review<br>(n=41, 52). | Patient perceptions about the screening tool<br>were not assessed, but clients were willing to<br>complete it, and five clients voluntarily gave<br>positive feedback. | Chart reviews show decreased documentation of a<br>reproductive plan (22% vs 6%, p=.02), and no change in<br>documentation of contraceptive counseling (20% vs.<br>13%, p=0.36) or the patient's contraceptive method<br>(20% vs. 37%, p=.08). |  | Chart reviews show no<br>significant change in<br>documentation current<br>contraceptive method<br>(20% vs. 37%, p=.08). |
| (Stulberg,<br>Dahlquist et al.<br>2019) | Cross-sectional<br>pre-post pilot<br>patient survey<br>(n=29, 34), no<br>control group. |  | The percentage of clients who said their provider<br>discussed birth control increased from 52% to 76%<br>(p=.04); percentage who recommended LARC increased<br>from 10% to 32% (p=.04). |  |  |
| **FPQ/RepLI** |  |  |  |  |  |
| (Baldwin,<br>Overcarsh et al.<br>2018) | RCT with post-test survey<br>(clients: OKQ<br>n=39, FPQ n=37;<br>providers OKQ<br>n=43, OKQ n=36). | FPQ patients were more likely than OKQ<br>patients to find the tool helpful and use it to<br>track reproductive health goals (76% vs. 51%,<br>p=.02). | FPQ clients were as likely as OKQ clients to find the tool<br>helpful in communicating their reproductive goals to<br>their provider (68% vs. 66%, p=.88).<br>FPQ providers were less likely than OKQ providers to<br>agree the tool helped focus their counseling, but the<br>effect was not significant (37% vs. 50%, p=.25). |  |  |
| (Madrigal,<br>Stempinski-Metoyer et al.<br>2019) | Post-test only<br>study with clients<br>(n=790) and<br>providers (n=66). | Completion of the FPQ/RepLI tool by a health<br>educator took about 5 minutes. 92% of<br>patients found the tool helpful and would use<br>it to track their reproductive goals.<br>Most providers agreed that the tool was<br>useful to facilitate discussing reproductive<br>health (91%) and that this type of tool is<br>needed (83%). | Most patients agreed the tool helped them think about<br>their personal goals (94%) and helped communicate<br>their personal goals to the provider (90%).<br>Most providers agreed that the tool helped them<br>understand the patient's reproductive plan (91%), help<br>focus their counseling (92%), and improved the family<br>planning counseling they provided (73%). |  |  |
| **Smart Choices** |  |  |  |  |  |
| (Koo, Wilson et<br>al. 2017) | Post-test only<br>study with<br>intervention<br>(n=126) and<br>control clients<br>(n=214). | The average completion time was 14 minutes. | In multivariate analyses, intervention women rated their<br>visit more patient-centered than controls (mean score<br>3.9 vs. 3.7, p<.05). Intervention women reporting<br>discussing more sexual health topics than control women<br>(1.2 vs. 0.9, p<.10). No effect was found on the number<br>of childbearing-related topics that were discussed. | After controls,<br>intervention women<br>knew 11.1<br>contraceptive<br>methods vs. 10.7 for<br>the control group<br>(p<.001). | After controls,<br>intervention women<br>were less likely than<br>controls to adopt IUDs or<br>implants (9% vs. 20%),<br>and more likely to select<br>oral contraceptives (64%<br>vs. 54%, p<.10). |
| **My Birth Control**<br>(Dehlendorf, Fitzpatrick et al. 2019) | RCT with post-test survey of providers (n=28) and clients (n=758). |  | Intervention clients reported higher interpersonal quality of counseling (OR=1.45 [1.03-2.05]) and greater satisfaction with side-effects information (OR=1.61 [1.11-2.33]). The tool had no effect on patient satisfaction with how the provider helped with method choice (OR=1.30 [.93-1.82]). | The tool improved knowledge of several contraceptive attributes. E.g., intervention clients were more likely to know that IUDs are more effective than pills (OR=2.65 [1.94-3.62]), that methods that cause period to stop are safe (OR=1.86 [1.28-2.71]), and that implants to not affect fertility (OR=1.54 [1.14-2.07]). | The tool had no effect on satisfaction with the chosen method (OR=1.19 [.88-1.61]), or on method continuation at 7 months (OR=.89 [.65-1.22]). |
| (Dehlendorf, Reed et al. 2019) | Qualitative assessment of providers (n=15). | All providers found it acceptable and feasible to incorporate the tool in their practice. Some noted that use of the tool prior to the visit sometimes slowed clinic flow. Most providers noted the tool was acceptable to clients but could be difficult for patients not used to the technology.<br>Use of the tool increased overall visit time by 12 minutes. | Nearly all providers reported the tool made contraceptive counseling more efficient and let them allocate more time to issues the patient wanted to discuss. | Providers reported the tool improved patient's pre-counseling knowledge of contraceptive options and method attributes. | Providers said the tool helped patients be more engaged and active in contraceptive method selection. |
| **My Path**<br>(Callegari, Nelson et al. 2021) | Cross-sectional pre-post pilot patient survey with intervention (n=30) and control group (n=28). | Most clients liked the tool, found it easy to understand and felt comfortable answering the questions. Average completion time was 11 minutes.<br>Providers did not think the tool increased their workload or hurt the clinic flow. | Most providers agreed it made counseling more efficient and helped them discuss pregnancy goals and contraceptives.<br>93% of intervention clients vs. 68% of control clients reported discussing pregnancy or contraceptive needs (p<.05). Scores for self-efficacy in communicating with providers improved more among intervention than control clients (0.8 vs. 0.2; p<.05). Use of the tool did not affect clients' rating of provider communication quality. | Scores for correct knowledge improved more among intervention than control clients (1.7 vs. 0.2; p<.01). | The tool had no significant effect on the likelihood of switching from non-prescription to prescription contraceptive methods. |
| **iMACC**<br>(Dev, Woods et al. 2019) | Qualitative assessment of clients (n=25) and providers (n=17). | Most clients and providers reported that iMACC was easy to use and self-explanatory; clients had no issues comprehending questions and material. Average completion time was 15 minutes.<br>Most providers said iMACC reduced their workload because it addresses common questions; a few worried that women's increased knowledge about contraceptives would take longer to counsel. | Clients valued the confidentiality of the tool and felt it would allow adolescents to answer more honestly.<br>Most providers noted the tool allowed women to ask more questions; some claimed most women had already decided on a method before meeting them. | iMACC helped clients understand their contraceptive options and potential side-effects and dispel myths. | iMACC made clients feel empowered to make informed decisions about methods most suitable for them. |

Our review indicates that all six tools were well accepted by both providers and clients. Clients generally liked the tools and found them easy to understand and use, but providers noted that patients with limited exposure to technology may have more difficulty using computer-based tools like My Birth Control (and hence also My Path). The tools are typically completed immediately before the provider visit, which takes about 5-15 minutes. Although providers had split opinions as to whether the tools affected the patient flow, this does not appear to be a major concern. Similarly, providers had little concern about the tools increasing their workload.

All tools also improved various aspects of quality of care. Consistent with the main focus of OKQ and FPQ/RepLI, the available evidence shows that 66-90% of clients reported that these tools helped them think about their reproductive goals and to communicate those goals to their provider (Baldwin, Overcarsh et al. 2018, Madrigal, Stempinski-Metoyer et al. 2019, Callegari, Nelson et al. 2021).

There is also some evidence that suggests that various tools can help providers to focus their counseling (Baldwin, Overcarsh et al. 2018, Dehlendorf, Reed et al. 2019, Madrigal, Stempinski-Metoyer et al. 2019, Callegari, Nelson et al. 2021). The proportion of providers who reported that the tool helped focuses their counseling was 50% for OKQ, 37% for FPQ alone, 92% for FPQ/RepLI. Studies of My Birth Control and iMACC indicated that such tools can make the counseling more efficient by enabling providers to spend more time on issues the patient wanted to discuss and by allowing clients to ask more questions (Dehlendorf, Reed et al. 2019, Dev, Woods et al. 2019).

In addition, the data suggest that decision-making tools can help make the provider-patient interaction more patient-centered, although some of the observed effects are relatively small. A pre-post analysis of Smart Choices found that the tool was associated with a small but statistically significant increase in perceived patient-centeredness, while a randomized controlled trial of My Birth Control found that the tool increased ratings of the perceived interpersonal quality of the counseling session. Use of Smart Choices, My Birth Control, My Path and iMACC all appear to have helped increase knowledge of contraceptive methods and their attributes (Koo, Wilson et al. 2017, Dehlendorf, Fitzpatrick et al. 2019, Dev, Woods et al. 2019, Callegari, Nelson et al. 2021).

Despite these increases in patient-centeredness of the interaction and the improved contraceptive knowledge, there is little evidence that these decision-making tools have improved family planning outcomes. For example, a randomized controlled trial of My Birth Control found no evidence that the tool improved satisfaction with the chosen method or method continuation after seven months (Dehlendorf, Fitzpatrick et al. 2019). Nevertheless, there are indications that decision-making tools can be invaluable for improving women’s contraceptive decision-making experience. Users of My Birth Control were 1.45 times (p=.03) more likely than the control group to give their visit the highest rating for the interpersonal quality of care. A qualitative analysis of women who used iMACC reported that the tool made them feel empowered to make informed decisions about which methods are best for them (Dev, Woods et al. 2019). A study of Smart Choices also found that use of the tool decreased adoption of long-acting methods from 20% to 9%, in favour of an increase in oral contraceptives from 54% to 64% (Koo, Wilson et al. 2017). Although this finding may seem counter-intuitive, it suggests that method effectiveness may not always be women’s main priority when selecting a contraceptive method, which confirms the importance of patient-centered care.

### Limitations of the review process and evidence

Our rapid review was limited to two commonly used databases, which implies that we may have missed other relevant articles. Because our review identified few studies that were conducted in developing countries, it is unclear to what extent our key findings can be generalized to family planning clients and providers in those regions. We restricted our review to publications in peer-reviewed journals and omitted the grey literature. While this restriction is likely to have yielded higher quality publications, it may have resulted in the exclusion of newer, lesser known approaches for patient-centered family planning counseling and support. Although there is ample literature on patient-centered family planning counseling and support, most of it focuses on support with the initial method choice. Our review identified few studies that address counseling and support for family planning customers after they have adopted their chosen method. Because tools to promote patient-centered family planning are in the early stages of development, several of the tools are relatively new. Hence, the evidence-base is still limited and often based on pilot studies with small samples.

## Discussion

Our systematic review aimed to assess how patient-centered family planning counseling has been conceptualized in the literature, and to identify tools that are being used to make counseling more patient-centered. We identified 33 peer-reviewed articles that discussed a strategy or intervention for providing patient-centered family planning counseling or provided evidence about the impact of such approaches.

Our findings show that there is no universally agreed upon definition of patient-centered family planning counseling. Only 18 of the 33 reviewed documents included a clear definition or described key domains of patient-centered family planning counseling. Consistent with Holt, Caglia et al. (2017), we found that although the definitions and terminology used tend to vary, the role of individual clients’ preferences, needs and values is widely recognized. Furthermore, it is accepted that respect for the patient and informed decision-making are essential.

Our review of how various studies have attempted to measure the level of patient-centeredness of the provider-patient interaction identified a wide range of measurement approaches, reflecting the lack of a universally accepted definition. However, we found that commonly used measurement questions can be grouped into six main areas: 1) soliciting information about the clients’ reproductive preferences; 2) soliciting the clients’ personal opinion about specific methods; 3) soliciting information about the expected role of the provider in the decision-making process; 4) the provider’s level of compassion and empathy; 5) the provider’s level of respect for the client; and 6) providing the patient opportunities to ask questions. Dehlendorf, Fox et al. (2021) have developed a Person-Centered Contraceptive Counseling (PCCC) scale that further condensed these topics to 1) respecting the patient as a person, 2) letting the patient say what matters to them about their birth control method, 3) taking the client’s preferences about birth control seriously, and 4) giving enough information to enable the patient to make the best contraceptive decision. Wider adoption of the PCCC scale in family planning surveys would generate consistency in measurement of patient-centered family planning counseling, which would strengthen the evidence base and may also help move the field toward a common definition of the concept.

The studies in our review provided detailed descriptions of six tools that aim to help make the interaction between family planning providers and their clients more patient-centered. Two of these tools focus on the clients’ reproductive goals, while the other four are better described as contraceptive decision-making tools. To better understand how these tools operate, we assessed which domains of patient-centered care each tool is trying to address. Consistent with Gagliardi, Nyhof et al. (2019), we used the six domains of patient-centered care identified by McCormack, Treiman et al. (2011).

Mapping of the tools against these domains revealed that none of the tools fully address all domains of patient-centered care. The domains most emphasized are “the exchange of information” and “facilitating decision-making”. All tools collect information about the client, which then guides the counseling, and four of the six tools are decision-making aids. Although “fostering the relationship between provider and client” is a quintessential domain of patient-centered care, none of the tools address it directly. However, several of the tools include provisions for the clients to list any questions or concerns they would like the provider to address, which may help address the power imbalance. To some extent, the four contraceptive decision-making tools facilitate “recognizing and responding to patient emotions” by inquiring how the patient feels about previously used contraceptive methods, specific method attributes, and potential partner influences. As expected, the four contraceptive decision-making tools identify and/or recommend contraceptive preferences that match the client’s needs and preferences. Generally speaking, the tools do little to directly address the domains “managing uncertainty” or “enabling self-management”, both of which are largely left to the discretion of the provider.

Our review included only very limited evidence about the impact of tools to promote patient-centered family planning counseling, and most of it stems from fairly small pilot studies. Nevertheless, several important findings emerged. Because providers are likely to be concerned that patient-centered counseling may be time-consuming and increase their workload, it is self-explanatory that provider acceptability of the tools is essential for them to be widely adopted. As noted by Baldwin, Overcarsh et al. (2018), “Given time constraints in clinics, any job aid needs to be easy to integrate and efficient, and should provide enough information to facilitate individualized counseling.” Our systematic review shows that all the tools were generally well accepted by both clients and providers. Because the tools are typically used immediately before the provider visit, their use does not normally increase the providers’ workload, and many providers indicated that the tools made their counseling more efficient. These findings are encouraging for the wider adoption of tools to promote patient-centered family planning counseling.

Some of the included studies found that use of contraceptive decision-making tools improved the clients’ perceptions about the quality of care and empowered them to make informed decisions about which contraceptive method is best suited for their own needs and preferences. However, while several studies show that use of the tools improved knowledge of contraceptive methods (including method features and/or side-effects), there is no evidence yet that the tools improved satisfaction with the client’s chosen contraceptive method or reduced method discontinuation. Hence, there is a need for further research to assess what can be done to translate the improved counseling experience generated by the decision-making tools into better family planning outcomes.

There is also a need to expand testing of these tools in low-income countries and other contexts, where they may be hard to use in their current form. Several studies showed that background characteristics, such as race, ethnicity, age, geographic location, sexual orientation, pregnancy norms, and pregnancy history, affect the acceptance and effectiveness of these family planning tools (Downey, Arteaga et al. 2017, Dev, Woods et al. 2019, Madrigal, Stempinski-Metoyer et al. 2019, Carvajal, Mudafort et al. 2020). Hence, tailoring the content of the tools to the specific cultural context may increase their effectiveness.

It is equally important to ensure that the mode of implementation of the tool is appropriate for the target population. For example, use of the tools may be hampered because the health facility does not have the required technology or because clients may not have the skills to use them (Dehlendorf, Reed et al. 2019). In numerous societies, open discussions about sexual and reproductive health are stigmatized. Hence, a lack of privacy in the waiting area may deter clients from using the tool. Because of such constraints, it will be important to adapt tools to promote patient-centered counseling to the local context. The current tools tend be computer or smart-phone based, which further limits their use in low-income settings. However, in many low-income countries large segments of the population have a feature phone (i.e., a phone with voice and text message capabilities, but only limited internet features). Feature phones are already being used extensively to access family planning information hotlines, including both operator-assisted hotlines and IVR (Interactive Voice Response) services. Feature phones also provide access to WhatsApp-based health information services, including family planning information. Given the widespread use of feature phones to access existing family planning information and services, there are important opportunities to adapt tools for patient-centered counseling for use with those services.

## Data Availability

The PRISMA checklists and underlying data are available at:

Harvard Dataverse: “Replication Data for: Patient-centered approaches for family planning counseling and support: A systematic review”, https://doi.org/10.7910/DVN/OV3M9Z.

## Consent

This study does not contain human subject data.

## Reporting guidelines

The article adheres to the PRISMA reporting guidelines for systematic reviews.

## Author Contributions

Meekers, D: Conceptualization, Formal Analysis, Writing – Original draft preparation; Elkins: Formal Analysis, Writing – Review and editing; Obozekhai: Writing – Review and editing.

## Data Availability

The PRISMA checklists and underlying data are available at Harvard Dataverse: "Replication Data for: Patient-centered approaches for family planning counseling and support: A systematic review", https://doi.org/10.7910/DVN/OV3M9Z.

https://doi.org/10.7910/DVN/OV3M9Z

## Acknowledgments

This systematic research was conducted to support the implementation of the “Rapidly test innovations to support contraceptive continuation” project, which is implemented in collaboration with DKT Nigeria and Data Scientists Network (DSN) in Nigeria.

## References

Assaf, S., W. Wang and L. Mallick (2017). “Quality of care in family planning services in Senegal and their outcomes.” BMC Health Serv Res 17(1): 346.

Baldwin, M. K., P. Overcarsh, A. Patel, L. Zimmerman and A. Edelman (2018). “Pregnancy intention screening tools: a randomized trial to assess perceived helpfulness with communication about reproductive goals.” Contracept Reprod Med 3: 21.

Brandi, K. and L. Fuentes (2020). “The history of tiered-effectiveness contraceptive counseling and the importance of patient-centered family planning care.” Am J Obstet Gynecol 222(4S): S873–S877.

Bruce, J. (1990). “Fundamental elements of the quality of care: a simple framework.” Stud Fam Plann 21(2): 61–91.

Callegari, L. S., K. M. Nelson, D. E. Arterburn, C. Dehlendorf, S. L. Magnusson, S. K. Benson, E. B. Schwarz and S. Borrero (2021). “Development and Pilot Testing of a Patient-Centered Web-Based Reproductive Decision Support Tool for Primary Care.” J Gen Intern Med 36(10): 2989–2999.

Callegari, L. S., E. V. Tartaglione, S. L. Magnusson, K. M. Nelson, D. E. Arteburn, J. Szarka, L. Zephyrin and S. Borrero (2019). “Understanding Women Veterans’ Family Planning Counseling Experiences and Preferences to Inform Patient-Centered Care.” Womens Health Issues 29(3): 283–289.

Carvajal, D. N., P. C. R. Mudafort, B. Barnet and A. E. Blank (2020). “Contraceptive Decision Making Among Latina Immigrants: Developing Theory-Based Survey Items.” Hisp Health Care Int 18(4): 181–190.

Dehlendorf, C., J. Fitzpatrick, E. Fox, K. Holt, E. Vittinghoff, R. Reed, M. P. Campora, A. Sokoloff and M. Kuppermann (2019). “Cluster randomized trial of a patient-centered contraceptive decision support tool, My Birth Control.” Am J Obstet Gynecol 220(6): 565 e561–565 e512.

Dehlendorf, C., E. Fox, I. A. Silverstein, A. Hoffman, M. P. Campora Perez, K. Holt, R. Reed and D. Hessler (2021). “Development of the Person-Centered Contraceptive Counseling scale (PCCC), a short form of the Interpersonal Quality of Family Planning care scale.” Contraception 103(5): 310–315.

Dehlendorf, C., J. T. Henderson, E. Vittinghoff, K. Grumbach, K. Levy, J. Schmittdiel, J. Lee, D. Schillinger and J. Steinauer (2016). “Association of the quality of interpersonal care during family planning counseling with contraceptive use.” Am J Obstet Gynecol 215(1): 78 e71–79.

Dehlendorf, C., J. T. Henderson, E. Vittinghoff, J. Steinauer and D. Hessler (2018). “Development of a patient-reported measure of the interpersonal quality of family planning care.” Contraception 97(1): 34–40.

Dehlendorf, C., R. Reed, J. Fitzpatrick, M. Kuppermann, J. Steinauer and K. Kimport (2019). “A mixed-methods study of provider perspectives on My Birth Control: a contraceptive decision support tool designed to facilitate shared decision making.” Contraception 100(5): 420–423.

Dev, R. (2023). iMacc Counseling Tool. Vancouver, Canada.

Dev, R., N. F. Woods, J. A. Unger, J. Kinuthia, D. Matemo, S. Farid, E. R. Begnel, P. Kohler and A. L. Drake (2019). “Acceptability, feasibility and utility of a Mobile health family planning decision aid for postpartum women in Kenya.” Reprod Health 16(1): 97.

Donnelly, K. Z., C. Dehlendorf, R. Reed, D. Agusti and R. Thompson (2019). “Adapting the Interpersonal Quality in Family Planning care scale to assess patient perspectives on abortion care.” J Patient Rep Outcomes 3(1): 3.

Donnelly, K. Z., T. C. Foster and R. Thompson (2014). ”What matters most? The content and concordance of patients’ and providers’ information priorities for contraceptive decision making.” Contraception 90(3): 280–287.

Downey, M. M., S. Arteaga, E. Villasenor and A. M. Gomez (2017). “More Than a Destination: Contraceptive Decision Making as a Journey.” Womens Health Issues 27(5): 539–545.

Fox, E., A. Reyna, N. M. Malcolm, R. B. Rosmarin, L. B. Zapata, B. N. Frederiksen, S. B. Moskosky and C. Dehlendorf (2018). “Client Preferences for Contraceptive Counseling: A Systematic Review.” Am J Prev Med 55(5): 691–702.

Gagliardi, A. R., B. B. Nyhof, S. Dunn, S. L. Grace, C. Green, D. E. Stewart and F. C. Wright (2019). “How is patient-centred care conceptualized in women’s health: a scoping review.” BMC Womens Health 19(1): 156.

Gawron, L. M., S. Simonsen, M. M. Millar, J. Lewis-Caporal, S. Patel and R. G. Simmons (2022). “Pregnancy Risk Screening and Counseling for Women Veterans: Piloting the One Key Question in the Veterans Healthcare Administration.” South Med J 114(3): 150–155.

Gomez, A. M. and M. Wapman (2017). “Under (implicit) pressure: young Black and Latina women’s perceptions of contraceptive care.” Contraception 96(4): 221–226.

Hamon, J. K., J. Hoyt, S. Krishnaratne, A. A. Barbra, J. Morukileng, N. Spilotros, M. Mbembe, S. Marcus and J. Webster (2022). “Perceptions of quality and the integrated delivery of family planning with childhood immunisation services in Kenya and Uganda.” PLoS One 17(6): e0269690.

Hazel, E., D. Mohan, E. Chirwa, M. Phiri, F. Kachale, P. Msukwa, J. Katz and M. A. Marx (2021). “Disrespectful care in family planning services among youth and adult simulated clients in public sector facilities in Malawi.” BMC Health Serv Res 21(1): 336.

Holt, K., J. M. Caglia, E. Peca, J. M. Sherry and A. Langer (2017). “A call for collaboration on respectful, person-centered health care in family planning and maternal health.” Reprod Health 14(1): 20.

Holt, K., I. Zavala, X. Quintero, D. Mendoza, M. C. McCormick, C. Dehlendorf, E. Lieberman and A. Langer (2018). “Women’s preferences for contraceptive counseling in Mexico: Results from a focus group study.” Reprod Health 15(1): 128.

Hunter, M. S., S. Meieran and H. Rosenau. (2012). ”One Key Question. Integrating Pregnancy Intention Screening.” Retrieved May 27, 2023, from https://www.chcs.org/media/OKQ-Webinar-618.pdf.

Institute of Medicine, C. o. Q. o. H. C. i. A. (2001). Crossing the Quality Chasm: A New Health System for the 21st Century. Washington (DC), National Academies Press.

Kamhawi, S., C. Underwood, H. Murad and B. Jabre (2013). “Client-centered counseling improves client satisfaction with family planning visits: evidence from Irbid, Jordan.” Glob Health Sci Pract 1(2): 180–192.

Koo, H. P., E. K. Wilson and A. M. Minnis (2017). “A Computerized Family Planning Counseling Aid: A Pilot Study Evaluation of Smart Choices.” Perspect Sex Reprod Health 49(1): 45–53.

Loyola Briceno, A. C., J. Kawatu, K. Saul, K. DeAngelis, B. Frederiksen, S. B. Moskosky and L. Gavin (2017). “From theory to application: using performance measures for contraceptive care in the Title X family planning program.” Contraception 96(3): 166–174.

Madrigal, J. M., K. Stempinski-Metoyer, A. E. McManus, L. Zimmerman and A. Patel (2019). “The family planning quotient and reproductive life index (FPQ/RepLI) tool: a solution for family planning, reproductive life planning and contraception counseling.” Reprod Health 16(1): 125.

McCormack, L. A., K. Treiman, D. Rupert, P. Williams-Piehota, E. Nadler, N. K. Arora, W. Lawrence and R. L. Street, Jr. (2011). “Measuring patient-centered communication in cancer care: a literature review and the development of a systematic approach.” Soc Sci Med 72(7): 1085–1095.

Meekers, D., A. Elkins and V. Obozekhai (2023). “Replication Data for: Patient-centered approaches for family planning counseling and support: A systematic review”, Harvard Dataverse.

Morse, J. E., S. Ramesh and A. Jackson (2017). “Reassessing Unintended Pregnancy: Toward a Patient-centered Approach to Family Planning.” Obstet Gynecol Clin North Am 44(1): 27–40.

Page, M. J., J. E. McKenzie, P. M. Bossuyt, I. Boutron, T. C. Hoffmann, C. D. Mulrow, L. Shamseer, J. M. Tetzlaff, E. A. Akl, S. E. Brennan, R. Chou, J. Glanville, J. M. Grimshaw, A. Hrobjartsson, M. M. Lalu, T. Li, E. W. Loder, E. Mayo-Wilson, S. McDonald, L. A. McGuinness, L. A. Stewart, J. Thomas, A. C. Tricco, V. A. Welch, P. Whiting and D. Moher (2021). ”The PRISMA 2020 statement: an updated guideline for reporting systematic reviews.” BMJ 372: n71.

Page, M. J., D. Moher, P. M. Bossuyt, I. Boutron, T. C. Hoffmann, C. D. Mulrow, L. Shamseer, J. M. Tetzlaff, E. A. Akl, S. E. Brennan, R. Chou, J. Glanville, J. M. Grimshaw, A. Hrobjartsson, M. M. Lalu, T. Li, E. W. Loder, E. Mayo-Wilson, S. McDonald, L. A. McGuinness, L. A. Stewart, J. Thomas, A. C. Tricco, V. A. Welch, P. Whiting and J. E. McKenzie (2021). “PRISMA 2020 explanation and elaboration: updated guidance and exemplars for reporting systematic reviews.” BMJ 372: n160.

Singal, S., S. K. Sikdar, S. Kaushik, P. Singh, N. Bhatt, G. Samandari, M. Pal, L. Cagatay, A. Arya and K. A. O’Connell (2021). ”Understanding factors associated with continuation of intrauterine device use in Gujarat and Rajasthan, India: a cross-sectional household study.” Sex Reprod Health Matters 29(2): 1–16.

Soin, K. S., P. T. Yeh, M. E. Gaffield, C. Ge and C. E. Kennedy (2022). “Health workers’ values and preferences regarding contraceptive methods globally: A systematic review.” Contraception 111: 61–70.

Stulberg, D. B., I. H. Dahlquist, J. Disterhoft, J. K. Bello and M. S. Hunter (2019). “Increase in Contraceptive Counseling by Primary Care Clinicians After Implementation of One Key Question(R) at an Urban Community Health Center.” Matern Child Health J 23(8): 996–1002.

Ti, A., R. Burns, E. S. Barnert, C. Sufrin and C. Dehlendorf (2019). “Perspectives on Patient-Centered Family Planning Care from Incarcerated Girls: A Qualitative Study.” J Pediatr Adolesc Gynecol 32(5): 491–498.

Welti, K., J. Manlove, J. Finocharo, B. Faccio and L. Kim (2022). “Women’s experiences with person-centered family planning care: Differences by sociodemographic characteristics.” Contracept X 4: 100081.

Wilson, E. K., K. E. Krieger, H. P. Koo, A. M. Minnis and K. Treiman (2014). “Feasibility and acceptability of a computer-based tool to improve contraceptive counseling.” Contraception 90(1): 72–78.

Worthington, R. O., J. Oyler, A. Pincavage, N. A. Baker, M. Saathoff and J. Rusiecki (2020). “A Novel Contraception Counseling and Shared Decision-Making Curriculum for Internal Medicine Residents.” MedEdPORTAL 16: 11046.

